# A multicenter, prospective, cross-sectional, genotype-phenotype and longitudinal natural history study of Andersen-Tawil syndrome

**DOI:** 10.1101/2022.05.26.22275429

**Authors:** Sanjeev Rajakulendran, Martin Tristani-Firouzi, Reza Sadjadi, James Cleland, Rabi Tawil, Giovanni Meola, Valeria Sansone, Jaya Trivedi, Stephen C Cannon, Michael G Hanna, Robert C. Griggs, the CINCH Investigators

## Abstract

**Objective:** A multi-center, prospective, cross-sectional natural history study to define the clinical phenotype of Andersen-Tawil syndrome, validate its current diagnostic criteria, explore genotype-phenotype correlations, and establish clinically relevant endpoints for use in therapeutic trials.

**Methods:** Participants were followed at yearly intervals for two years. Outcome measures included attack frequency and duration, neurophysiological exercise testing and interictal muscle strength. Cardiac endpoints were QTc interval, presence of U-waves, frequency of ventricular ectopy and arrhythmias. Participants completed the SF-36 and underwent *KCNJ2* gene analysis.

**Results:** 28 participants were enrolled. The age range was 17 to 82 years. 23 participants harbored mutations in *KCNJ2*, including a new mutation, Y68D. All exhibited at least one skeletal feature with 26/28 exhibiting two or more. Common physical abnormalities were a small mandible (89%), low set ears (82%) and micromelia of hands or feet (71%). 26 participants reported periodic paralysis. The frequency of attacks varied from 12/week to 1/year, and duration from 12 minutes to 21 days. Common triggers for attacks were prolonged rest (85%) and exercise/exertion (73%). 20/25 had an abnormal long exercise test. A prolonged QTc interval was identified in 36% participants, U waves in 39% and ambulatory ECGs demonstrated runs of ventricular tachycardia in 32% and more than 10 000 ventricular couples in 14% of participants.

**Interpretation:** There is extensive heterogeneity in both the spectrum and severity of Andersen-Tawil syndrome. Overall, the symptoms result in a significant impairment in quality of life. The cardiac and neurophysiological data could serve as outcome measures in future treatment trials.

## INTRODUCTION

In 1971, Andersen and colleagues reported a child who exhibited “intermittent muscle weakness, cardiac extra-systoles and developmental anomalies” ^1^ in what was the first description of the triad of clinical features that would later characterise Andersen-Tawil syndrome (ATS), an autosomal dominant, multi-system ion channel disorder ^2-4^. The intermittent muscle weakness or periodic paralysis may last from hours to days, is typically precipitated by prolonged rest or rest following exertion and is usually ‘hypokalemic’ although normal and raised potassium levels during an attack may occur. Permanent interictal weakness is common^5^. Cardiac manifestations include prominent U waves, prolonged QTc, frequent premature ventricular complexes (PVC), non-sustained polymorphic or bidirectional ventricular tachycardia (VT), and dilated cardiomyopathy ^6-12^, with the potential for life-threatening events^13^, and sudden cardiac death ^14^. In some, a cardiac phenotype may be the sole manifestation of ATS^15-17^. The spectrum of skeletal anomalies includes micrognathia, low set ears, short stature, clinodactyly, syndactyly and micromelia, valvular heart disease, structural kidney abnormalities, epilepsy, and cognitive impairment have also been reported in association with ATS^18-20^

Mutations in *KCNJ2*, which encodes the inwardly rectifying potassium channel Kir2.1, account for approximately 70% of individuals with a clinical phenotype of ATS, with locus heterogeneity likely to account for the remainder ^4,19,21,22^. Kir2.1 is widely expressed in the brain, skeletal and cardiac muscle ^23,24^. The normal function of this channel is to stabilize the resting membrane potential of the cell and shape the terminal portion of the action potential^4^. Most mutations reduce Kir2.1 current by a dominant-negative mechanism ^6,25,26^, with impairment of PIP_2_ binding and diminished trafficking of channels to the cell surface also implicated in the pathogenesis^21,27^. Although the loss of Kir2.1 channel function is understood to account for attacks of paralysis and cardiac dysrhythmias^28-30^, the mechanisms underlying the developmental skeletal abnormalities remain unknown.

ATS is the least studied of the skeletal muscle channelopathies. However, the nature of its multisystem involvement poses interesting questions about the diverse biological roles of Kir2.1, especially with respect to skeletal development. Our aims were to obtain prospective multi-modal data to define the clinical spectrum of ATS, explore genotype-phenotype correlations, consider the impact of symptoms on quality of life and establish outcome measures for use in future interventional trials.

## MATERIALS AND METHODS

### Study Design

This study was a multi-center, prospective, cross-sectional longitudinal study of Andersen-Tawil syndrome as part of the NIH-funded Rare Disease Clinical Research Network’s Consortium for Clinical Investigation of Neurological Channelopathies (CINCH). Participants were recruited from seven sites in the USA, one in the UK, one in Canada, and one in Italy. The evaluators were trained to perform all outcomes in a standardized manner at investigator meetings. Informed consent was obtained from all study participants, and the protocol was approved by the institutional review boards or ethic committees at all participating sites (supplementary table 3). Table 1 sets out the inclusion criteria.

**Table 1.** Inclusion criteria for the study.

| Inclusion Criteria |  |
| --- | --- |
| **I** *Individuals with clinically confirmed diagnosis of ATS as defined by at least two of the following three features:* |  |
|  | **1 Muscle features**<br>A) Presence of clear-cut episodes of transient muscle weakness with or without a fixed deficit, typically following exertion or prolonged rest<br>*OR*<br>B) An atypical history with otherwise typical exam findings (absent reflexes with normal sensation during an episode<br>*OR*<br>C) Unexplained hypokalaemia between episodes<br>*OR*<br>D) An abnormal long-exercise nerve conduction study |
|  | **2 Cardiac features**<br>A) Prolonged QTc interval on 12-lead ECG according to standard criteria<br>*OR*<br>B) Ventricular ectopy including uniform or multifocal PVCs, polymorphic VT, or bidirectional VT |
|  | **3 Skeletal features**<br>A) Presence of two or more physical features: low set ears; hypertelorism; small mandible; clinodactyly; syndactyly; micromelia of hands or feet |
|  | **OR** Individuals with one of the three above criteria with at least one other affected family member meeting two criteria |
|  | **OR** Individuals not meeting clinical criteria but possessing the KCNJ2 mutation |
| **II** *Male and female participants age 10 and older meeting the criteria above who are able to comply with the study conditions* |  |

### Patient evaluation

At their baseline visit, participants completed a standardised questionnaire covering the following domains: Periodic paralysis history, cardiac symptoms, medical history, family history, and medications history. Baseline data included age, gender, self-reported race and ethnicity, and educational history. All participants underwent a standardised physical examination.

### Laboratory tests and genetic analysis

The following blood tests were undertaken at baseline: Potassium, urea, creatinine, thyroid stimulating hormone and creatine kinase. For those participants who had not had genetic analysis prior to enrolment, consent was obtained and their DNA was sequenced for mutations in *KCNJ2* at the University of Rochester using a standard method described previously.

### Cardiac investigations

A digital 12-lead ECG was obtained at baseline and at annual follow-up visits. QTc prolongation was defined as ≥ 470 ms. Twenty-four-hour ambulatory ECG monitoring was obtained at baseline for all participants. A subset of subjects underwent high-resolution 24-hour continuous 12-lead digital ECG monitoring (1000 Hz sampling rate using H12+ Holter monitors, Mortara Instruments, Milwaukee, WI). Raw 24-hour Mortara H12+ recordings were analyzed using in-house software as described previously ^31^. Lead II QT interval was measured using the conventional tangent method. U and T wave amplitudes were measured from the RMS signal^31^. All values were averaged over a 10 min epoch.

### MMT / QMT

Muscle strength was tested for the following: shoulder abduction, elbow flexion/extension, wrist flexion/extension, hip flexion/ extension/abduction, knee flexion/extension, ankle plantar flexion/dorsiflexion. A total manual muscle score was calculated by adding up individual muscle scores^32^. Quantitative hand grip dynamometry was obtained using a force transducer connected to automatic capturing software (QMA system, Computer Source). Each hand grip recorded was the best of three maximal voluntary isometric contractions recorded in kg force.

### Interactive voice response system for documentation of attacks of weakness

Participants were asked to use an automated interactive voice response system to report every attack for eight consecutive weeks after recruitment. This system assessed symptom presence (yes/no) during attacks, trigger for the attack, the date and the duration of the attack. Individuals were asked to estimate the attack severity and the effect on daily functioning on a three-point scale (mild, moderate or severe). Any further episodes on that day could also be reported. This real time voice response diary was adopted to reduce recall bias that can occur with paper diary entries^33,34^.

### Quality of life assessment

Quality of life was assessed using SF-36 version 2 at baseline and annually. This is a self-reported questionnaire grouped into 8 quality of life domains and two summary components that generates scores pertaining to physical and mental health wellbeing that can be compared to normalized control data, with an average of 50 and a standard deviation of 10^35^.

### Neurophysiology

At each visit participants underwent a McManis short exercise test as described by Fournier and colleagues, and a prolonged exercise test using a standardized protocol^36-39^. CMAPs were recorded for 5 minutes pre-exercise, during 5 minutes of exercise and then for 50 minutes post-exercise. In addition, needle EMG of proximal and distal muscles was performed. Post-exercise compound muscle action potential amplitudes were calculated as percent change from the average pre-exercise baseline measurement. The hand was placed in a neutral position and temperature was monitored throughout. Abnormal decrement was determined as 20% in the prolonged exercise test, as previously reported^37^.

## RESULTS

### General characteristics

A total of 28 participants fulfilled the eligibility criteria for the study and were recruited. Fifteen were male. The age range was between 17 and 82 years (median of 41 years; mean of 42 years). All participants were of Northern White European ethnicity. With respect to educational status, eleven of the 28 obtained a bachelor’s degree or equivalent and four left formal education without the equivalent of a high school diploma. The average length of formal education for the cohort was 14 years.

Periodic paralysis was the presenting symptom in 24/28 (85%) of participants and cardiac symptoms in 3/28. Nineteen (68%) reported a positive family history. Assessment of psychiatric co-morbidity revealed depression in 9/28 (32%) and anxiety in 5/28 (18%); 9/28 (32%) experienced migraines; 2/28 (7%) had epilepsy and 4/26 (15%) reported learning difficulties.

Thyroid function was normal in all tested individuals. Prior to the onset of the study, records from three individuals (participants 2, 19 and 25) revealed a low interictal potassium level (2.6 mmol/L, 3.0 mmol/L and 3.0mmol/L respectively).

### Physical characteristics

Examination of physical characteristics revealed that mandibular hypoplasia (25/28; 89%), low set years (23/28, 82%) micromelia of hands or feet (20/28, 71%), clinodactyly (18/28, 64%), a high arched palate (18/28; 64%) and hypertelorism (14/28; 50%) were the most common physical findings in our cohort (table 2 1). A broad nose (8/28; 28%), scoliosis (8/28; 28%) and syndactyly (7/28; 25%) were less common. Eight individuals (28%) had ptosis on examination at some point. The average male height was 168.7 cm (range, 159-178 cm) and for a female was 153.5 cm (range 132-163). Only one individual was observed to have muscle atrophy on inspection at baseline (Supplementary table 1); atrophy was reported in another individual at 12 months follow-up; 2/28 were reported to have facial weakness at the baseline visit, an additional participant at 12 months follow-up and a fourth participant at 24 months having exhibited normal facial strength at baseline and 12 months. The average head circumference for males was 56.1 cm and 54.5 for females. Examination of eye movements was normal in all participants at baseline with the exception of participant 4, whose horizontal ocular movements were restricted.

**Table 2.**
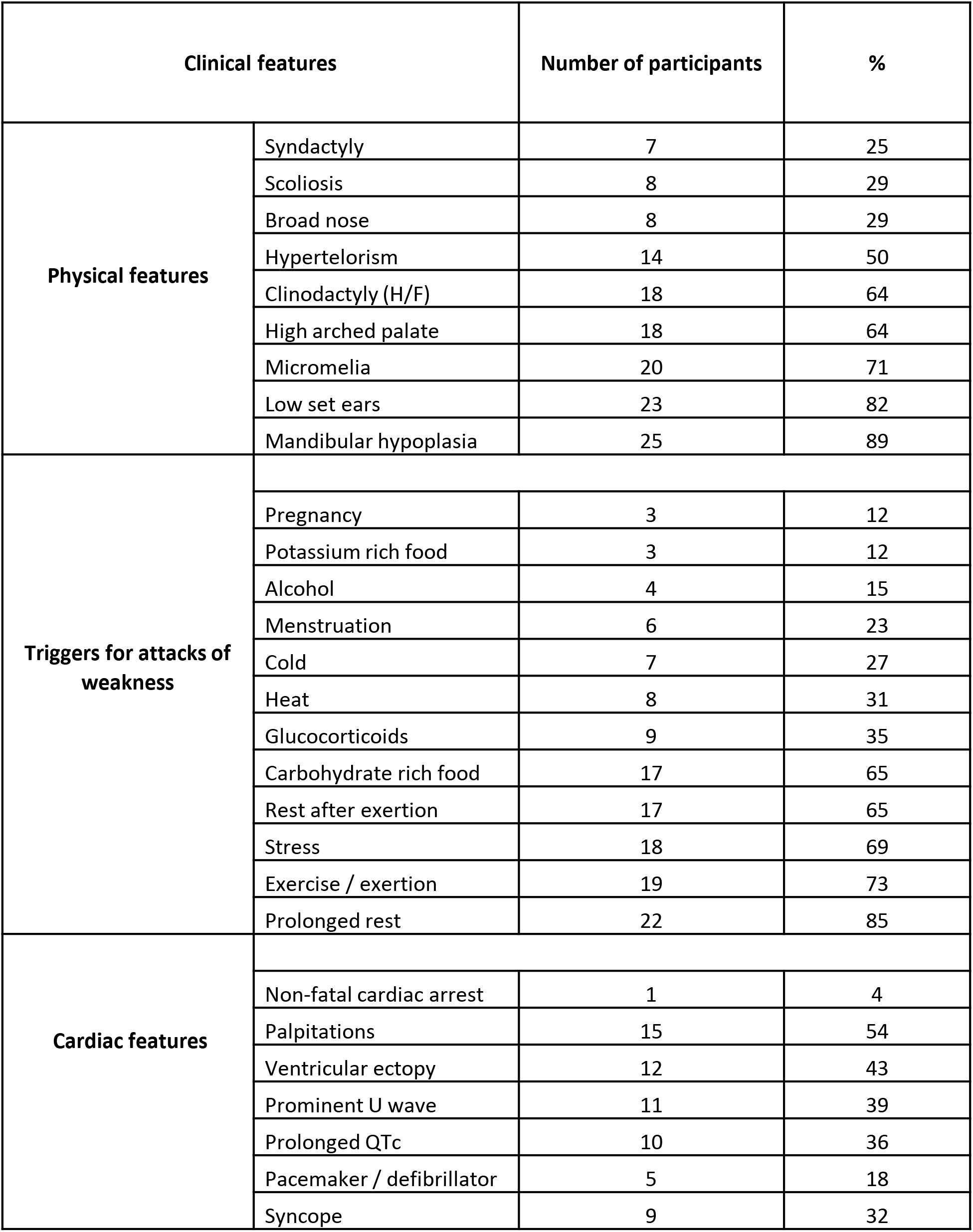
Clinical features in the 28 participants.

| Clinical features |  | Number of participants | % |
| --- | --- | --- | --- |
| Physical features | Syndactyly | 7 | 25 |
|  | Scoliosis | 8 | 29 |
|  | Broad nose | 8 | 29 |
|  | Hypertelorism | 14 | 50 |
|  | Clinodactyly (H/F) | 18 | 64 |
|  | High arched palate | 18 | 64 |
|  | Micromelia | 20 | 71 |
|  | Low set ears | 23 | 82 |
|  | Mandibular hypoplasia | 25 | 89 |
| Triggers for attacks of weakness |  |  |  |
|  | Pregnancy | 3 | 12 |
|  | Potassium rich food | 3 | 12 |
|  | Alcohol | 4 | 15 |
|  | Menstruation | 6 | 23 |
|  | Cold | 7 | 27 |
|  | Heat | 8 | 31 |
|  | Glucocorticoids | 9 | 35 |
|  | Carbohydrate rich food | 17 | 65 |
|  | Rest after exertion | 17 | 65 |
|  | Stress | 18 | 69 |
|  | Exercise / exertion | 19 | 73 |
|  | Prolonged rest | 22 | 85 |
| Cardiac features |  |  |  |
|  | Non-fatal cardiac arrest | 1 | 4 |
|  | Palpitations | 15 | 54 |
|  | Ventricular ectopy | 12 | 43 |
|  | Prominent U wave | 11 | 39 |
|  | Prolonged QTc | 10 | 36 |
|  | Pacemaker / defibrillator | 5 | 18 |
|  | Syncope | 9 | 32 |

### Periodic Paralysis

A total of 26/28 (93%) participants experienced episodes of periodic paralysis which in all but two manifested as bilateral extremity weakness. However, these two individuals were only seen at baseline visit. 17/26 (65%) reported permanent interictal muscle weakness at some point in the study. The range for the age of onset of attacks was 1–50 years. Twenty-two participants experienced muscle pain / tenderness.

The Modified Rankin score was used to assess the average degree of disability arising from muscle symptoms. 5/26 (19%) reported ‘no disability’; 12/26 (46%) reported ‘minor disability’ which led to restriction of lifestyle but do not interfere with capacity for self-care; 9/26 (35%) reported ‘moderate disability’ which significantly interfered with lifestyle or prevented totally independence. 6/26 reported progression in disability over 1or 2 years either from ‘no-disability’ to ‘minor disability’ or ‘minor disability’ to ‘moderate disability’

There was variability in the frequency of attacks, ranging from one per year (participant 12; R67W mutation) to 12 per week (participant 5; R312C mutation). Of the 26 participants who experienced episodes of paralysis, 11 reported a frequency of at least once a week (range 1–12/week), 11 reported a frequency of less than once a week (range 1-3 month); one reported a frequency of once a year (participant 12; R67W); one reported an attack every two months and the attack frequency was not recorded in two participants.

Twelve out of 13 who had a trial of acetazolamide reported subjective benefit (table 3). Four participants reported improvement with potassium supplementation, one with amiloride and one with dichlorphenamide (data not shown).

**Table 3.** Muscle features of all 28 participants, including genetic results and response to acetazolamide.

| Participant ID | Genetics | Permanent weakness | Attack duration (Longest) | Attack duration (shortest) | Attack Frequency | Acetazolamide outcome |
| --- | --- | --- | --- | --- | --- | --- |
| 1 | R67W | Yes | 71 | 1 | 2/week |  |
| 2 | R82W | Yes | 192 | 1 | 5/week | Improvement |
| 3 | Negative | No | 48 | 24 | 1/month |  |
| 4 | R218W | Yes | 240 | 72 | 1/month | Improvement |
| 5 | R312C | Yes | 4 | 0.5 | 12/week | Improvement |
| 6 | R218W | Yes | 504 | 24 | 1/month |  |
| 7 | Y68D | Yes | 336 | 72 | 1/month | Improvement |
| 8 | Y68D | Yes | 72 | 12 | 2/month | Improvement |
| 9 | Negative | Yes | 2 | 0.2 | 5/week | Improvement |
| 10 | L217P | No | 168 | 72 | 1/month | Improvement |
| 11 | R67W | Yes | 2 | 1 | 1/two months |  |
| 12 | R67W | Yes | 336 | 2 | 1/ year | Improvement |
| 13 | R312C | No | 72 | 1 | 4/week | Improvement |
| 14 | Negative | Yes | 3 | 3 | 3/week |  |
| 15 | Y68D | Yes | 504 | 12 | 3/month | Improvement |
| 16 | Y68D | No | 48 | 4 | 1/week |  |
| 17 | Negative | N/A | N/A | N/A | N/A |  |
| 18 | T75M | Yes | N/A | N/A | N/A |  |
| 19 | R218W | Yes | 12 | 0.2 | 1/week |  |
| 20 | Y82D | Yes | 96 | 4 | 1/week | Improvement |
| 21 | V302M | Yes | 48 | 1 | 1/week |  |
| 22 | Negative | Yes | 72 | 2 | 1/week |  |
| 23 | R67W | No | 72 | 3 | 1/month |  |
| 24 | N216H | No | 5 | 0.5 | 1/month |  |
| 25 | N216H | No | 6 | 3 |  |  |
| 26 | R82Q | No | 20 | 2 | 1/month |  |
| 27 | N216H | No | 2 | 1 | 1/month | Improvement |
| 28 | N216H | Yes | 120 |  |  |  |

The most common triggers for attacks of paralysis (table 2) were prolonged rest (22/26, 85%) exercise/exertion (19/26, 73%) and stress (18/26, 69%). 20/26 cited dietary triggers with the consumption of carbohydrate rich food (17/26, 65%) the most common reason. In contrast, the consumption of alcohol (4/26, 15%), potassium rich food (3/26, 11.5%) and temperature (cold 7/26, 26.9%; heat 8/26, 30.8%) were less likely to precipitate attacks of paralysis. Three individuals reported an increase in frequency of attacks during pregnancy. Other triggers reported included sleep deprivation (2/26), emotional excitement (2/26), infection (1/26), caffeine (1/26), high salt intake (1/26) and use of vibrating tools (1/26).

The majority of participants experienced attacks upon awakening (21/26, 80.8%) and 12 (57.1%) participants reported attacks in the evening; 12 (57.1%) reported no diurnal pattern to the attacks.

### Neurophysiology

25 participants underwent long exercise test at baseline visit. 15/25 had greater than 50% and 20/25 participants had greater than 20% reduction in the area; 5 patients had greater than 50% and 20/25 patients had greater than 20% reduction in the amplitude (Figure 1). 5/25 had less than 20% drop in the area or amplitude. Therefore, 80% of the participants had an abnormal test.

**Figure 1.**
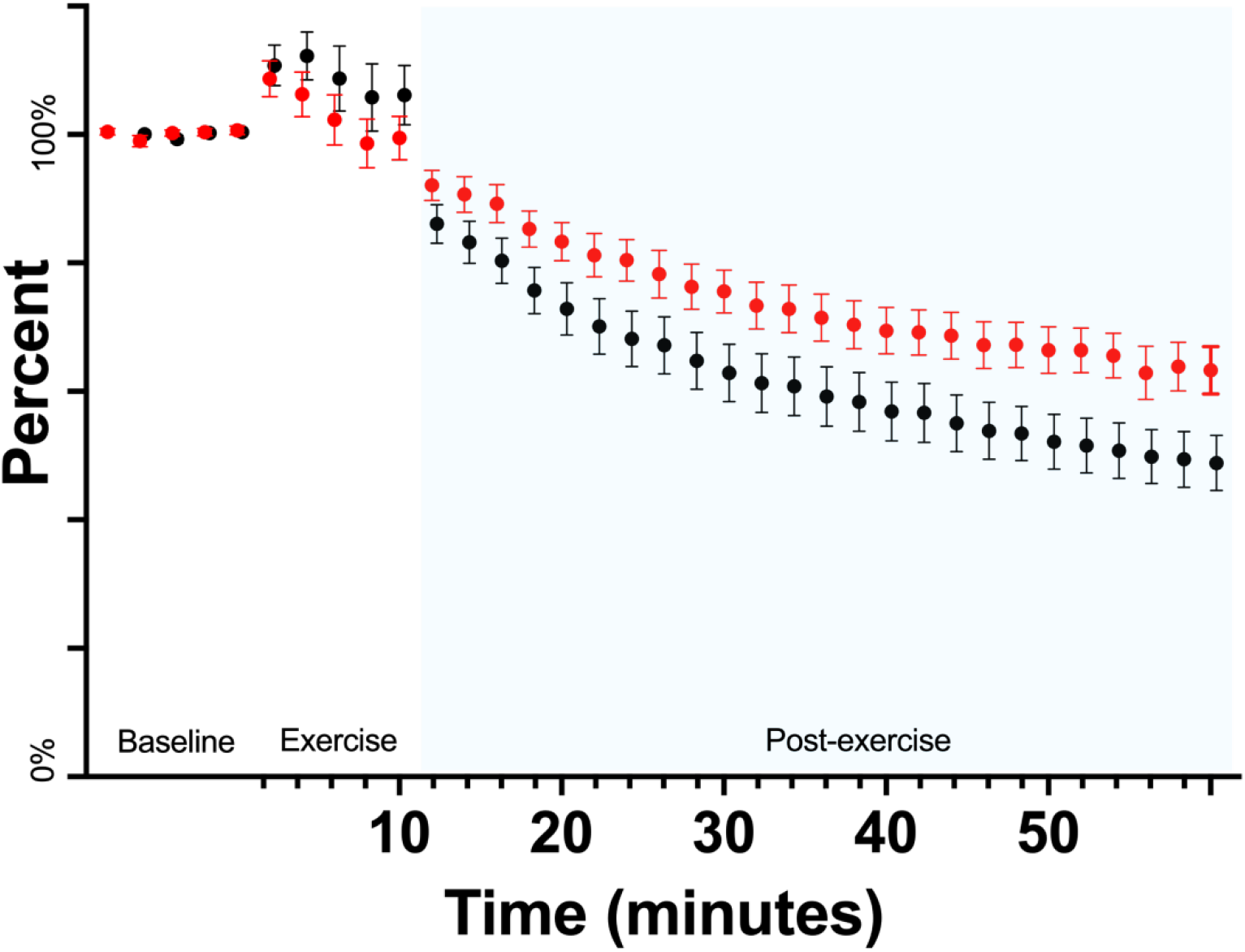
Long exercise test, patients show increase in compound motor action potential (CMAP) amplitude (black bars) and area (red bars) during the exercise followed by drop post-exercise

### MMT / QMT

MMT and QMT scores are summarized in table 4 and visually shown in figure 2. Most patients had overall normal strength in MMT testing. Participant 4 and 25 were noted to have diminished muscle bulk and mild distal weakness (on MMT scores and Grip dynamometry). Only two patients were noted to have facial weakness (participant 4 and 13). 5/28 participants had ptosis throughout study visits. Deep tendon reflexes were diminished in 8/28 patients. A single patient had brisk deep tendon reflexes without identifiable aetiology. Mean grip dynamometry was 26.1 95% CI [22.8;29.5] KgF on the left and 28.2 95% CI [24.9;31.5] KgF on the right. There was no definite proximal or distal pattern of weakness. Additionally, there was no significant difference between MMT and QMT scores between visits. There was no significant difference between right and left sided muscles.

**Table 4.**
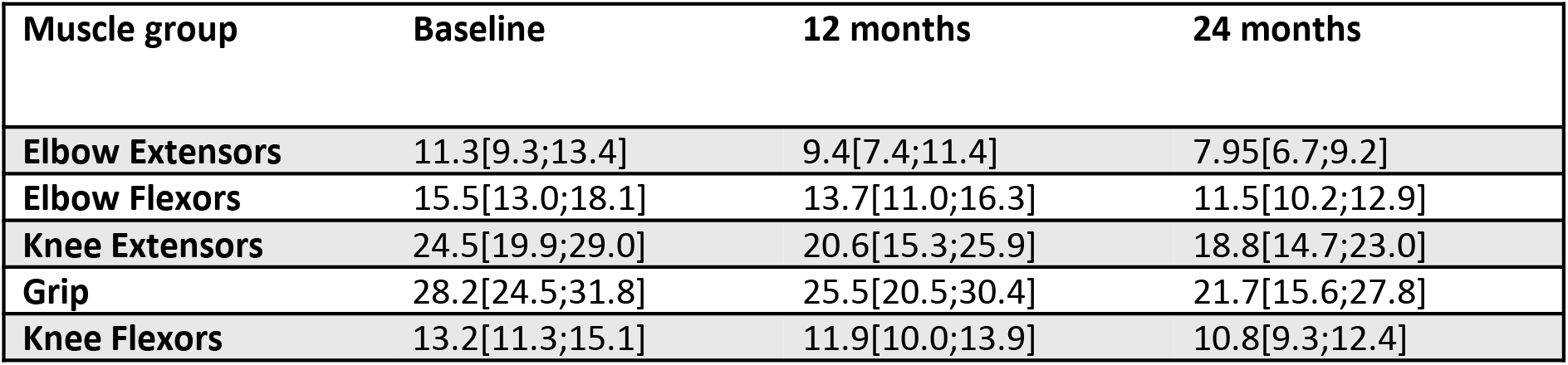
Average right and left quantitative muscle strength; mean[95%CI] in KgF

**Figure 2.**
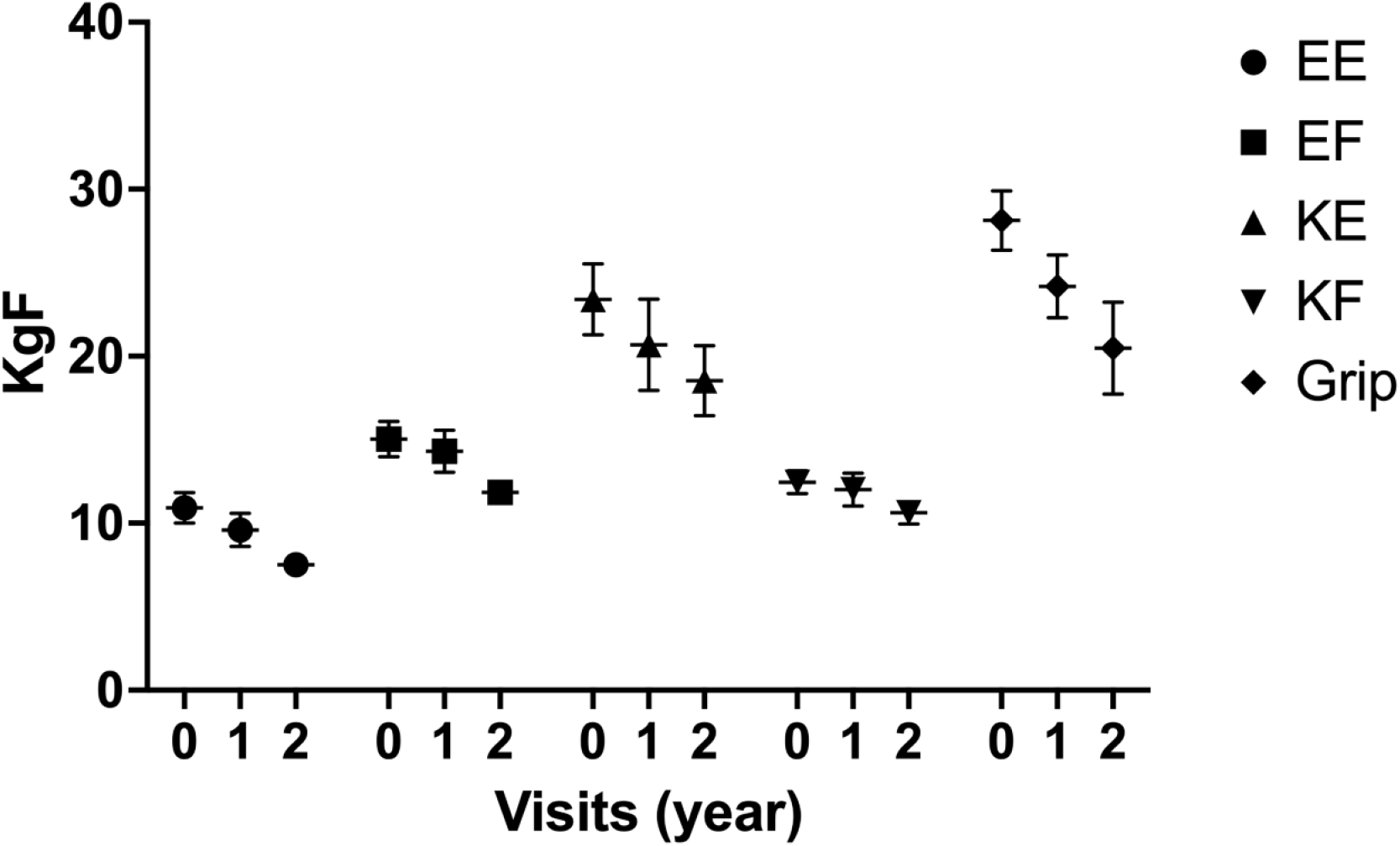
Average right and left quantitative muscle strength; mean [95%CI] in KgF. EE= elbow extension, EF= elbow flexion, KE=knee extension, KF=knee flexion

### Cardiac features

Nineteen participants (68%) reported cardiac symptoms with 15 (54%) experiencing palpitations and 9 (32%) syncope. The frequency of palpitations varied: daily in 4/15; weekly in 3/15; monthly in 3/15 and rarely in 5/15. Syncope was a ‘rare’ occurrence in nine participants. Five individuals (18%) had either a pacemaker or defibrillator *in situ* of which three were negative for mutations in *KCNJ2*; one of these had suffered a non-fatal cardiac arrest.

A total of 10/28 (36%) had prolonged corrected QT (QTc) interval on resting 12 lead ECG, defined as ≥ 470 ms. 11/28 (39%) had prominent U waves. Fourteen subjects underwent high-resolution Holter monitor recordings, allowing for sequential QT and U wave analyses. In 9 subjects the mean amplitude of the U wave exceeded 30% of the T wave amplitude (supplemental figure 1). The mean QTc values of 6 participants exceeded 470 ms (supplemental figure 2) over the 24-hour recording period.

There was considerable variability with respect to the frequency of ventricular ectopic beats, with 4 subjects experiencing more than 10,000 ectopic beats (figure 3; 24-hour ECG data from 28 participants). Ventricular couplets were detected in 13 subjects (46%). Nine had runs of ventricular tachycardia, 4 more than 5 runs and two in excess of 40 (one participant having 60 runs of VT).

**Figure 3.**
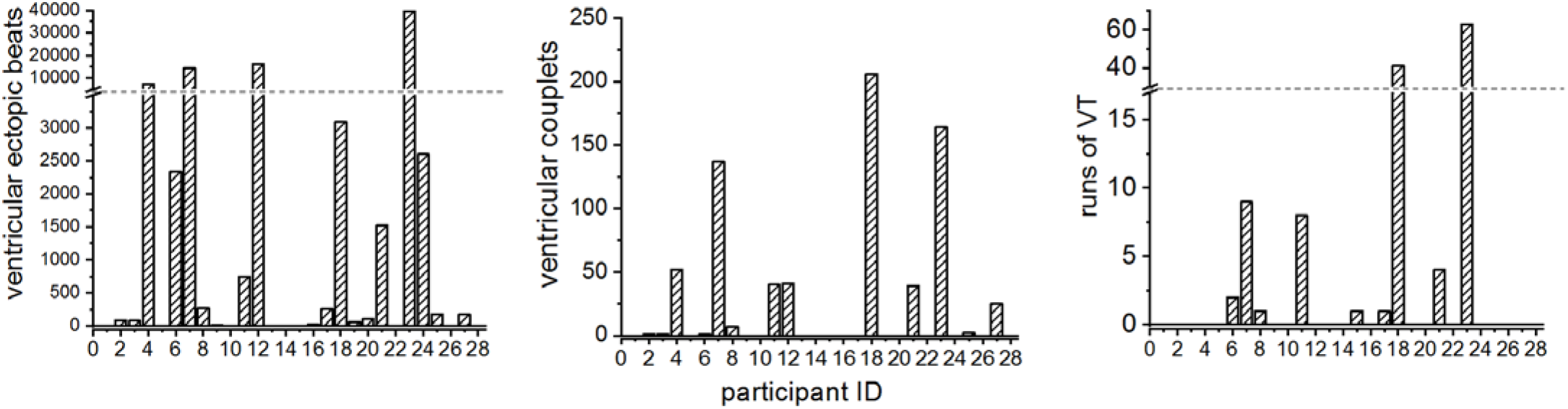
Cardiac outcomes. Ventricular ectopic beats, ventricular couplets and runs of VT (ventricular tachycardia) in the participants.

Stress and exertion/exercise were the most common triggers for cardiac symptoms (12/19, 63%) with 7/19 (37%) reporting carbohydrate loading, 6/19 (32%) rest after exertion and 4/19 (21%) prolonged rest as precipitants (Supplementary Figure 3).

### Quality of life

Twenty participants completed the SF-36 version 2 questionnaire at baseline, fifteen at 1 year and seven at 2 years (figure 4). At baseline, when compared to norm-based average scores with a mean of 50 and standard deviation of 10, all 8 domains were below normal with ‘Physical Function’ and ‘Role Physical’ (role limitations due to physical problems) the lowest with scores of 37.4 and 38.4 respectively, both of which were below one standard deviation of normal. Overall, higher scores were recorded in the mental subscales compared with physical subscale as reflected in the standardised summary scores (38.4 versus 48.5). There was a trend towards a decline in scores on the physical subscales at 1 year follow up with a significant reduction in ‘Physical Function’.

**Figure 4.**
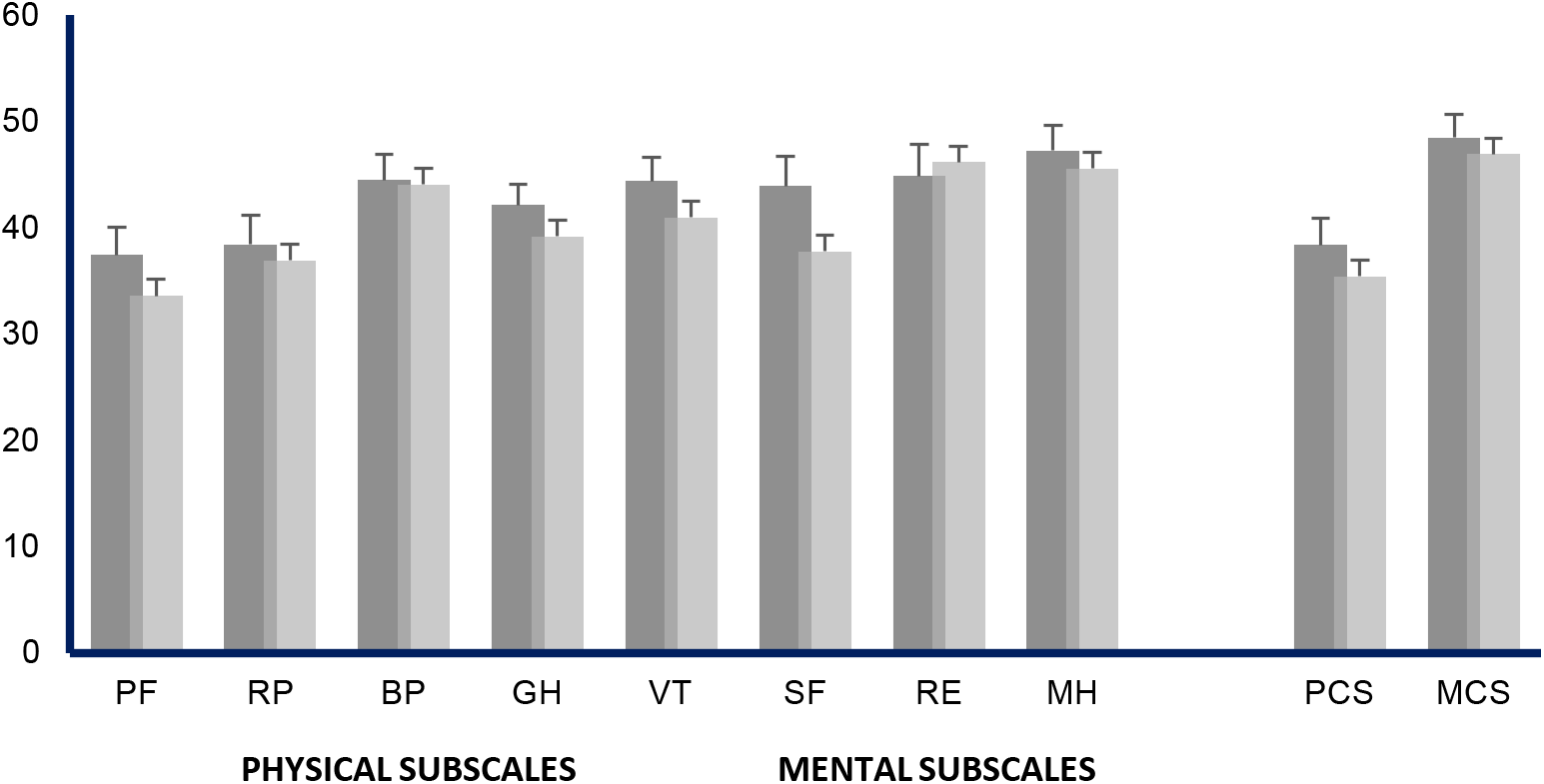
SF-36 domain scores in the assessment of quality of life. Mean = 50, standard deviation = 10. PF, physical function; RP, role physical; BP, bodily pain; GH, general health; VT, vitality; SF, social function; RE, role emotional; MH, mental health; PC, physical component; MC, mental component.

### Genotype-Phenotype correlation

Twenty-three individuals harbored mutations in *KCNJ2* (tables 3 and 5). Ten different missense mutations accounted for the genetic basis of ATS in these individuals. R67W, Y68D and N216H were the most common, each accounted for 4 individuals.

**Table 5.** Genotype-phenotype correlation

| Participant ID | KCNJ2 Genetics | Transient weakness | Abnormal long exercise test | Prolonged QT | Prominent U wave | Ventricular ectopy | 2 or more physical features | FH with 2 or more features |
| --- | --- | --- | --- | --- | --- | --- | --- | --- |
| 1 | R67W | Yes | Yes | No | Yes | No | Yes | Yes |
| 2 | R82W | Yes | Yes | Yes | Yes | Yes | Yes | No |
| 3 | Negative | Yes | No | Yes | No | Yes | Yes | No |
| 4 | R218W | Yes | Yes | Yes | Yes | Yes | Yes | Yes |
| 5 | R312C | Yes | No | Yes | No | No | Yes | Yes |
| 6 | R218W | Yes | Yes | No | No | No | Yes | Yes |
| 7 | Y68D | Yes | Yes | Yes | No | No | Yes | Yes |
| 8 | Y68D | Yes | Yes | Yes | No | No | Yes | Yes |
| 9 | Negative | Yes | Yes | No | No | Yes | Yes | Yes |
| 10 | L217P | Yes | No | Yes | No | No | Yes | Yes |
| 11 | R67W | Yes | Yes | Yes | No | Yes | Yes | Yes |
| 12 | R67W | Yes | Yes | Yes | No | Yes | Yes | Yes |
| 13 | R312C | Yes | No | No | No | No | Yes | Yes |
| 14 | Negative | Yes | No | Yes | Yes | No | Yes | No |
| 15 | Y68D | Yes | Yes | No | No | No | Yes | Yes |
| 16 | Y68D | Yes | Yes | No | No | No | Yes | Yes |
| 17 | Negative | No | Yes | Yes | Yes | Yes | Yes | Yes |
| 18 | T75M | No | No | No | Yes | Yes | Yes | Yes |
| 19 | R218W | Yes | Yes | Yes | No | Yes | Yes | No |
| 20 | Y82D | Yes | Yes | Yes | No | No | Yes | Yes |
| 21 | V302M | Yes | Yes | Yes | Yes | Yes | Yes | No |
| 22 | Negative | Yes | Yes | No | No | No | Yes | No |
| 23 | R67W | Yes | No | Yes | No | Yes | Yes | Yes |
| 24 | N216H | Yes | No | Yes | Yes | No | Yes | Yes |
| 25 | N216H | Yes | No | Yes | Yes | No | Yes | Yes |
| 26 | R82Q | Yes | No | Yes | Yes | No | Yes | No |
| 27 | N216H | Yes | No | Yes | Yes | No | Yes | Yes |
| 28 | N216H | Yes | No | Yes | No | Yes | Yes | Yes |

All participants regardless of genotype exhibited at least two distinctive skeletal features. R218W was associated with a more severe phenotype with the three affected participants exhibiting 8, 6 and 6 physical features. Two of these individuals also reported long duration of attacks (range 72-240 and 24-504 hours). However, there was discordance with the cardiac phenotype with electrocardiography of one participant demonstrating a prolonged QT interval, u waves and ventricular ectopy and another participant lacking all three features.

Although 26/28 participants reported episodes of transient weakness, there was considerable variability of the phenotype even in participants who harbored the same mutations with respect to the severity and frequency of periodic paralysis; one participant with R67W had two attacks a week whereas another with the same mutation reported one attack a year with some attacks lasting 336 hours whereas yet another with the identical variant described typical attacks of 1-2 hours duration. The R312C mutation in two participants was associated with frequent attacks (12/week and 4/week) and neither had abnormal long exercise tests.

The missense Y68D mutation has not been previously reported in the medical literature. It results in a substitution of Tyrosine for Aspartic acid at amino acid position 68 within the N terminus. All four participants had mandibular hypoplasia, low set ears, micromelia and clinodactyly. All four exhibited transient muscle weakness; two had prolonged attacks of paralysis (72-336 and 12-504 hours) and the frequency varied from 1/month to 1/week. All four had abnormal long exercise test results. On the whole, the cardiac phenotype was less prominent in these participants; none of the four had prominent u waves and infact two had normal ECGs; two had prolonged QT intervals.

Five participants did not harbor mutations in *KCNJ2*. The clinical, cardiac and neurophysiological findings are shown in supplementary table 2. Further genetic testing (e.g for *KCNJ5* mutations) was not undertaken. There was phenotypic heterogeneity with respect to the number of distinctive / skeletal / developmental features, varying from two to seven. All five experienced episodes of paralysis and all four who were tested had positive long exercise tests and three reported subjective benefit with acetazolamide. Four had typical electrocardiographic features and of note, three had either a pacemaker of defibrillator; one had suffered a cardiac arrest.

## DISCUSSION

This is the largest prospective study of Andersen-Tawil syndrome to date, encompassing all features of the disorder. Its prospective, longitudinal and multi-axial design, incorporating both ‘*KCNJ2*-positive’ and ‘mutation-negative’ cases, studied over a two-year period has enabled the capture of both quantitative and qualitative clinic data on this rare ion channel disorder. Our findings expand the phenotypic and genotypic spectrum of ATS, contribute important data about quality of life and ‘real-world’ information through the interactive voice response system, highlight the importance of diagnostic awareness and accuracy, and suggest important avenues of future research.

Twenty-three participants (82%) harbored mutations in *KCNJ2* including the new Y68D variant. Five participants had a typical phenotype for ATS but were ‘mutation-negative’, implying locus heterogeneity, a common feature among the voltage-gated channelopathies. Recently a mutation in *KCNJ5*, which encodes a G-protein-activated inwardly rectifying potassium channel 4 (Kir3.4) was identified in a *KCNJ2*-negative individual with flaccid paralysis and U-waves in whom there was no skeletal or dysmorphic features^22^; its prevalence however in ‘*KCNJ2*-negative’ cases is unknown. It is likely that the 30% of *KCNJ2*-negative cases reflect mutations in different genes, whose identification will be facilitated by the discovery of more ATS kindreds and the growing awareness of this disorder amongst neurologists and cardiologists. Close to 70% had a positive family history which merits discussion during consultations because of the potential for cardiac complications; and the possibility of other affected members in the family.

The number of mutation-negative cases was too small to undertake a definitive statistical comparison, but a qualitative assessment between this cohort and those who harbored *KCNJ2* mutations would suggest that the phenotype is indistinguishable with four out of the five mutation-negative participants exhibiting all three classical features of ATS. One finding of note is that three in the mutation-negative group had either a pacemaker or defibrillator *in situ*, supporting a cardiac assessment in all individuals and the need for both the identification of more *KCNJ2*-negative cases and research into their genetic basis.

Both cardiac symptoms and electrocardiographic abnormalities, including typical ‘signatures’ such as u-waves, prolonged QT intervals and bidirectional VT, are common in affected individuals; recognizing them is paramount in light of the potential for malignant cardiac dysrhythmias. This study is the first to systematically explore and confirm triggers for cardiac symptoms, and this finding will have clinical implications. Some affected individuals will present to cardiologists, either because of an isolated or predominant cardiac phenotype or because the muscle symptoms are minor or have not yet manifested. Awareness of the classical triad of ATS is therefore important within the cardiology sphere. Moreover, the recent observation of significant risk for life-threatening cardiac events in ATS^13^, further underscores the importance of a thorough cardiac evaluation in those in whom ATS is either confirmed or suspected.

Distinct skeletal abnormalities and characteristic facies are a core feature of ATS, particularly in those harbouring *KCNJ2* mutations. All 28 cases had at least two physical anomalies with mandibular hypoplasia, low set ears and micromelia predominating; clinodactyly, a high arched palate and hypertelorism were identified in over 50% of cases. Therefore, the conjunction of episodic muscle weakness with distinctive features is an important clue to the diagnosis and the latter should be actively examined for in all cases of periodic paralysis. Short stature was common and present in all twelve females. Ptosis, previously undescribed was identified in eight individuals.

In addition to the physical features, muscle symptoms were very common with the majority of participants reported episodes of transient weakness of varying onset (the latest onset was 50 years), frequency, duration and disability. Muscle pain and tenderness were widely reported. Despite the subjective reporting of permanent muscle weakness, there was no indication of muscle atrophy after a year, although clearly a longer follow-up interval would be required to ascertain whether muscle loss is a feature.

15% reported intellectual difficulties. Previous reports of individuals with ATS have highlighted mild learning difficulties with deficits in executive functioning and abstract reasoning^18^. Although duration of and achievements in education are helpful surrogates, we suggest that formal and detailed neuropsychometric testing should be undertaken on affected individuals to ascertain the specific pattern of involvement and further delineate the neurocognitive phenotype, as we suspect the figure is probably higher than what is reported in this study.

Two ‘mutation positive’ participants had epilepsy. Although this finding is not surprising, the figure is perhaps lower than expected in light of the strong association between mutations in members of the voltage-gated ion channel superfamily (sodium, potassium and calcium channels are all associated with epileptic phenotypes) and monogenic epilepsies and also taking into account the ‘extensive’ pathological spectrum already associated with *KCNJ2* in particular. To date, a potential link with seizures has only been described in one study^40^ in which afebrile seizures occurring in infancy were reported in 4/23 individuals from a Japanese cohort with genetically confirmed ATS. Epilepsy might be under-recognized and under-diagnosed in individuals with ATS.

Our data indicate that 11 participants reported a benefit with acetazolamide, supporting previous evidence for the efficacy of this medication in ATS. However, quantification of this improvement in terms of attack frequency and duration along with longitudinal data on retention rate and long-term benefit was not ascertained. Moreover, the subjectively reported benefits were retrospectively ascertained, suggesting the need for a prospective randomized trial, perhaps also examining the potential role for concomitant potassium supplementation. Not all individuals with transient muscle weakness had tried acetazolamide perhaps due to a lack of awareness or under-reporting of muscle symptoms. Dichlorphenamide and amiloride were also tried in participants, although the numbers were too small to draw any firm conclusions; they may have a role in those individuals who either do not tolerate or respond to acetazolamide

Many participants reported impaired quality of life, particularly on the physical subscale of the SF-36 and there was a trend toward deteriorating scores at the yearly follow-up interval. Although ATS is characterized by transient muscle weakness, many reported subjectively permanent weakness which affected their day-to-day physical roles. These symptoms may explain the relatively high incidence of depression in our cohort. Whether mood disorders are reactive or endogenous to ATS does merit further exploration^41^.

Although this study was able to recruit a large series of 28 participants, the drop out was significant by two years which impacted on specific outcomes measures such as muscle and cardiac symptoms and signs, quality of life and response to medications. In addition to supporting the need for a larger trial, we would also recommend a longer follow-up period.

## Data Availability

All data produced in the present study are available upon reasonable request to the authors

## Acknowledgements

The CINCH investigators thank the participants and referring clinicians at all sites.

## Author contributions

RCG, RNT, LP, SCC, MGH contributed to the conception and design of the study. MTF, RS, JC, RNT, GM, VS, JT, SCC, and SR contributed to the acquisition and analysis of data. MTF, RS, RCG and SR contributed to drafting the text and preparing the figures.

---

**Supplementary Table 1**.

Additional clinical features of all 28 participants. EOM, extraocular movements; N, normal; Ab, abnormal (weak); U, unknown; B, baseline, 12, 12 months review; 24, 24 months review.

**Supplementary Table 2**.

Clinical features of the KCNJ2 negative cohort, including physical characteristics, electrocardiography data and result of long exercise test.

**Supplementary Table 3**.

List of CINCH (Consortium for Clinical Investigation of Neurological Channelopathies) investigators.

**Supplementary Figure 1**

Mean amplitude of the U wave for all 28 participants over 24-hour recording period.

**Supplementary Figure 2**

Mean QTc values for all 28 participants over 24-hour recording period.

## REFERENCES

1. Andersen ED, Krasilnikoff PA, Overvad H. Intermittent muscular weakness, extrasystoles, and multiple developmental anomalies. A new syndrome? Acta Paediatr Scand 1971; 60(5): 559–64.

2. Sansone V, Griggs RC, Meola G, et al. Andersen’s syndrome: a distinct periodic paralysis. Ann Neurol 1997; 42(3): 305–12.

3. Tawil R, Ptacek LJ, Pavlakis SG, et al. Andersen’s syndrome: potassium-sensitive periodic paralysis, ventricular ectopy, and dysmorphic features. Ann Neurol 1994; 35(3): 326–30.

4. Plaster NM, Tawil R, Tristani-Firouzi M, et al. Mutations in Kir2.1 cause the developmental and episodic electrical phenotypes of Andersen’s syndrome. Cell 2001; 105(4): 511–9.

5. Child ND, Cleland JC, Roxburgh RH. Andersen-Tawil syndrome presenting as a fixed myopathy. Muscle Nerve 2013.

6. Tristani-Firouzi M, Jensen JL, Donaldson MR, et al. Functional and clinical characterization of KCNJ2 mutations associated with LQT7 (Andersen syndrome). J Clin Invest 2002; 110(3): 381–8.

7. Tristani-Firouzi M. Polymorphic ventricular tachycardia associated with mutations in KCNJ2. Heart Rhythm 2004; 1(2): 242–3.

8. Zhang L, Benson DW, Tristani-Firouzi M, et al. Electrocardiographic features in Andersen-Tawil syndrome patients with KCNJ2 mutations: characteristic T-U-wave patterns predict the KCNJ2 genotype. Circulation 2005; 111(21): 2720–6.

9. Deo M, Ruan Y, Pandit SV, et al. KCNJ2 mutation in short QT syndrome 3 results in atrial fibrillation and ventricular proarrhythmia. Proc Natl Acad Sci U S A 2013; 110(11): 4291–6.

10. Delannoy E, Sacher F, Maury P, et al. Cardiac characteristics and long-term outcome in Andersen-Tawil syndrome patients related to KCNJ2 mutation. Europace 2013.

11. Schoonderwoerd BA, Wiesfeld AC, Wilde AA, et al. A family with Andersen-Tawil syndrome and dilated cardiomyopathy. Heart Rhythm 2006; 3(11): 1346–50.

12. Rezazadeh S, Guo J, Duff HJ, Ferrier RA, Gerull B. Reversible Dilated Cardiomyopathy Caused by a High Burden of Ventricular Arrhythmias in Andersen-Tawil Syndrome. Can J Cardiol 2016; 32(12): 1576 e15–e18.

13. Mazzanti A, Guz D, Trancuccio A, et al. Natural History and Risk Stratification in Andersen-Tawil Syndrome Type 1. J Am Coll Cardiol 2020; 75(15): 1772–84.

14. Peters S, Schulze-Bahr E, Etheridge SP, Tristani-Firouzi M. Sudden cardiac death in Andersen-Tawil syndrome. Europace 2007; 9(3): 162–6.

15. Kimura H, Zhou J, Kawamura M, et al. Phenotype variability in patients carrying KCNJ2 mutations. Circ Cardiovasc Genet 2012; 5(3): 344–53.

16. Lopes CM, Zhang H, Rohacs T, Jin T, Yang J, Logothetis DE. Alterations in conserved Kir channel-PIP2 interactions underlie channelopathies. Neuron 2002; 34(6): 933–44.

17. Limberg MM, Zumhagen S, Netter MF, et al. Non dominant-negative KCNJ2 gene mutations leading to Andersen-Tawil syndrome with an isolated cardiac phenotype. Basic Res Cardiol 2013; 108(3): 353.

18. Yoon G, Quitania L, Kramer JH, Fu YH, Miller BL, Ptacek LJ. Andersen-Tawil syndrome: definition of a neurocognitive phenotype. Neurology 2006; 66(11): 1703–10.

19. Andelfinger G, Tapper AR, Welch RC, Vanoye CG, George AL, Jr., Benson DW. KCNJ2 mutation results in Andersen syndrome with sex-specific cardiac and skeletal muscle phenotypes. Am J Hum Genet 2002; 71(3): 663–8.

20. Davies NP, Imbrici P, Fialho D, et al. Andersen-Tawil syndrome: new potassium channel mutations and possible phenotypic variation. Neurology 2005; 65(7): 1083–9.

21. Donaldson MR, Jensen JL, Tristani-Firouzi M, et al. PIP2 binding residues of Kir2.1 are common targets of mutations causing Andersen syndrome. Neurology 2003; 60(11): 1811–6.

22. Kokunai Y, Nakata T, Furuta M, et al. A Kir3.4 mutation causes Andersen-Tawil syndrome by an inhibitory effect on Kir2.1. Neurology 2014; 82(12): 1058–64.

23. Raab-Graham KF, Radeke CM, Vandenberg CA. Molecular cloning and expression of a human heart inward rectifier potassium channel. Neuroreport 1994; 5(18): 2501–5.

24. Karschin C, Dissmann E, Stuhmer W, Karschin A. IRK(1-3) and GIRK(1-4) inwardly rectifying K+ channel mRNAs are differentially expressed in the adult rat brain. J Neurosci 1996; 16(11): 3559–70.

25. Ai T, Fujiwara Y, Tsuji K, et al. Novel KCNJ2 mutation in familial periodic paralysis with ventricular dysrhythmia. Circulation 2002; 105(22): 2592–4.

26. Hosaka Y, Hanawa H, Washizuka T, et al. Function, subcellular localization and assembly of a novel mutation of KCNJ2 in Andersen’s syndrome. J Mol Cell Cardiol 2003; 35(4): 409–15.

27. Ballester LY, Benson DW, Wong B, et al. Trafficking-competent and trafficking-defective KCNJ2 mutations in Andersen syndrome. Hum Mutat 2006; 27(4): 388.

28. Sung RJ, Wu SN, Wu JS, Chang HD, Luo CH. Electrophysiological mechanisms of ventricular arrhythmias in relation to Andersen-Tawil syndrome under conditions of reduced IK1: a simulation study. Am J Physiol Heart Circ Physiol 2006; 291(6): H2597–605.

29. Tsuboi M, Antzelevitch C. Cellular basis for electrocardiographic and arrhythmic manifestations of Andersen-Tawil syndrome (LQT7). Heart Rhythm 2006; 3(3): 328–35.

30. Bendahhou S, Donaldson MR, Plaster NM, Tristani-Firouzi M, Fu YH, Ptacek LJ. Defective potassium channel Kir2.1 trafficking underlies Andersen-Tawil syndrome. J Biol Chem 2003; 278(51): 51779–85.

31. Lux RL, Sower CT, Allen N, Etheridge SP, Tristani-Firouzi M, Saarel EV. The application of root mean square electrocardiography (RMS ECG) for the detection of acquired and congenital long QT syndrome. PLoS One 2014; 9(1): e85689.

32. Personius KE, Pandya S, King WM, Tawil R, McDermott MP. Facioscapulohumeral dystrophy natural history study: standardization of testing procedures and reliability of measurements. The FSH DY Group. Phys Ther 1994; 74(3): 253–63.

33. Palmblad M, Tiplady B. Electronic diaries and questionnaires: designing user interfaces that are easy for all patients to use. Qual Life Res 2004; 13(7): 1199–207.

34. Gwaltney CJ, Shields AL, Shiffman S. Equivalence of electronic and paper-and-pencil administration of patient-reported outcome measures: a meta-analytic review. Value Health 2008; 11(2): 322–33.

35. McHorney CA, Ware JE, Jr., Raczek AE. The MOS 36-Item Short-Form Health Survey (SF-36): II. Psychometric and clinical tests of validity in measuring physical and mental health constructs. Med Care 1993; 31(3): 247–63.

36. McManis PG, Lambert EH, Daube JR. The exercise test in periodic paralysis. Muscle Nerve 1986; 9(8): 704–10.

37. Fournier E, Arzel M, Sternberg D, et al. Electromyography guides toward subgroups of mutations in muscle channelopathies. Ann Neurol 2004; 56(5): 650–61.

38. Fournier E, Viala K, Gervais H, et al. Cold extends electromyography distinction between ion channel mutations causing myotonia. Ann Neurol 2006; 60(3): 356–65.

39. Tan SV, Matthews E, Barber M, et al. Refined exercise testing can aid DNA-based diagnosis in muscle channelopathies. Ann Neurol 2011; 69(2): 328–40.

40. Haruna Y, Kobori A, Makiyama T, et al. Genotype-phenotype correlations of KCNJ2 mutations in Japanese patients with Andersen-Tawil syndrome. Hum Mutat 2007; 28(2): 208.

41. Chan HF, Chen ML, Su JJ, Ko LC, Lin CH, Wu RM. A novel neuropsychiatric phenotype of KCNJ2 mutation in one Taiwanese family with Andersen-Tawil syndrome. J Hum Genet 2010; 55(3): 186–8.

