## Supplemental Figures and Tables for "A multicenter, prospective, cross-sectional, genotype-phenotype and longitudinal natural history study of Andersen-Tawil syndrome"

### Supplementary Table 1

| General physical examination | | | | Muscle examination | | | | | | | | | | | | | | |
|---|---|---|---|---|---|---|---|---|---|---|---|---|---|---|---|---|---|---|
| | | Height | Head circumference | Muscle bulk | | | Facial weakness | | | EOM | | | Ptosis | | | Absent/Reduced reflexes | | |
| ID | Sex | | | B | 12 | 24 | B | 12 | 24 | B | 12 | 24 | B | 12 | 24 | B | 12 | 24 |
| 1 | M | 168 | 55 | N | N | | N | Ab | | N | N | | Absent | Present | | N | N | |
| 2 | M | 174 | 57 | N | N | | N | N | | N | N | | Absent | Absent | | N | N | |
| 3 | F | U | 54 | N | | | N | | | N | | | Absent | | | N | | |
| 4 | F | 132 | 53 | Atrophy | Atrophy | Atrophy | Ab | Ab | N | Ab | Ab | Ab | Present | Present | Present | N | N | N |
| 5 | F | 158 | 54 | N | N | | N | N | | N | | | Absent | Absent | | N | N | |
| 6 | M | 172 | 57 | N | N | N | N | N | Ab | N | N | | Absent | Absent | Present | Reduced | Absent | N |
| 7 | M | 178 | 56 | N | N | N | N | N | N | N | N | N | Absent | Absent | Present | N | N | N |
| 8 | F | 156 | 56 | N | N | N | N | N | N | N | N | N | Present | Absent | Absent | Reduced | Reduced | Reduced |
| 9 | F | 163 | 54 | N | N | | N | N | | N | N | | Absent | Absent | | | N | |
| 10 | F | 149 | 58 | N | N | | N | N | | N | N | | Absent | Present | | Absent | N | |
| 11 | F | 160 | 54 | N | N | N | N | N | N | N | N | N | Present | Present | Present | N | | N |
| 12 | M | 170 | 55 | N | | N | N | N | N | N | N | N | Present | Absent | Absent | Reduced | Reduced | Absent |
| 13 | M | 180 | 60 | N | N | N | N | N | Ab | N | N | N | Absent | Absent | Present | N | N | N |
| 14 | F | 160 | U | N | N | | Ab | | | N | N | | Present | Absent | | N | N | |
| 15 | M | 160 | 54 | N | | N | N | N | | N | N | | Absent | Absent | | Reduced | Reduced | |
| 16 | M | 162 | U | N | | | n | N | N | N | N | N | Absent | Absent | Absent | Reduced | Reduced | Reduced |
| 17 | F | 150 | 56 | N | | | N | | | N | | | Absent | | | Increased | | |
| 18 | F | 151 | 53 | N | | | N | | | N | | | Absent | | | Reduced | | |
| 19 | F | 159 | 53 | N | N | | N | N | | N | N | | Absent | Absent | | Reduced | N | |
| 20 | M | 159 | U | N | N | | N | N | | N | N | | Absent | Absent | | N | N | |
| 21 | M | 172 | 55 | N | N | | N | N | | N | N | | Absent | Absent | | N | N | |
| 22 | M | 172 | 58 | N | | | N | | | N | | | Absent | | | N | | |
| 23 | M | 163 | 56 | N | N | | N | N | | N | N | | Absent | Absent | | N | N | |
| 24 | M | 160 | 55 | N | | | N | N | | N | N | | Absent | Absent | | N | N | |
| 25 | F | 151 | 60 | N | A | | N | N | | N | N | | Absent | Absent | | N | | |
| 26 | M | 170 | 58 | N | N | | N | N | | N | N | | Absent | Absent | | N | Reduced | |
| 27 | F | U | 49 | N | | | N | | | N | | | Absent | | | N | | |
| 28 | M | 171 | 53 | N | | | N | | | N | | | Absent | | | N | | |

### Supplementary Table 2

| | KCNJ2-negative participants | | | | |
|---|---|---|---|---|---|
| | Participant 3 | Participant 9 | Participant 14 | Participant 17 | Participant 22 |
| Sex | F | F | F | F | M |
| Short Stature | NA | No | No | Yes | No |
| Mandibular hypoplasia | Yes | Yes | No | Yes | Yes |
| Low-set years | Yes | Yes | No | No | Yes |
| Hypertelorism | Yes | No | No | No | Yes |
| High-arched palate | No | No | Yes | Yes | Yes |
| Micromelia | No | No | No | Yes | Yes |
| Clinodactyly | Yes | No | No | Yes | Yes |
| Syndactyly | No | No | No | No | No |
| Scoliosis | No | No | No | No | Yes |
| Prominent U wave | No | No | Yes | Yes | No |
| Prolonged QTc interval | Yes | No | Yes | Yes | No |
| Pacemaker | Yes | No | Yes | Yes | No |
| Non-fatal cardiac arrest | No | No | No | Yes | No |
| Episodes of periodic paralysis | Yes | Yes | Yes | Yes | Yes |
| Positive long exercise test | NA | Yes | Yes | Yes | Yes |

### Supplementary Table 3

| CINCH Investigators | Affiliation |
|---|---|
| Anthony A. Amato | Harvard / Brigham and Women's Hospital |
| Richard J. Barohn | University of South Florida |
| Brian N. Bundy | University of South Florida |
| Stephen C Cannon | University of Texas Southwestern |
| Emma Ciafaloni | University of Rochester |
| James Cleland | University of Rochester |
| Liz Dewar | MRC Centre for Neuromuscular Disease, UCL Institute of Neurology |
| Joseph Gomes | University of South Florida |
| Robert C. Griggs | University of Rochester |
| Michael G. Hanna | MRC Centre for Neuromuscular Disease, UCL Institute of Neurology |
| Kimberly Hart | University of Rochester |
| Barbara Herr | University of Rochester |
| Laura Herbelin | University of Kansas Medical Centre |
| Dipa Jayaseelan | MRC Centre for Neuromuscular Disease, UCL Institute of Neurology |
| Jeffrey Krischer | University of South Florida |
| Giovanni Meola | University of Milan |
| Louis Ptacek | University of California, San Francisco |
| Bonnie Patterson | University of South Florida |
| Sanjeev Rajakulendran | MRC Centre for Neuromuscular Disease, UCL Institute of Neurology |
| Valeria Sansone | University of Milan |
| Reza Seyedsadjadi | Harvard / Brigham and Women's Hospital |
| Rabi N. Tawil | University of Rochester |
| Martin Tristani-Firouzi | University of Utah |
| Veronica Tan | MRC Centre for Neuromuscular Disease, UCL Institute of Neurology |
| Jaya Trivedi | University of Texas Southwestern |
| Shannon Venance | University of Western Ontario |

**Supplementary Figure 1**

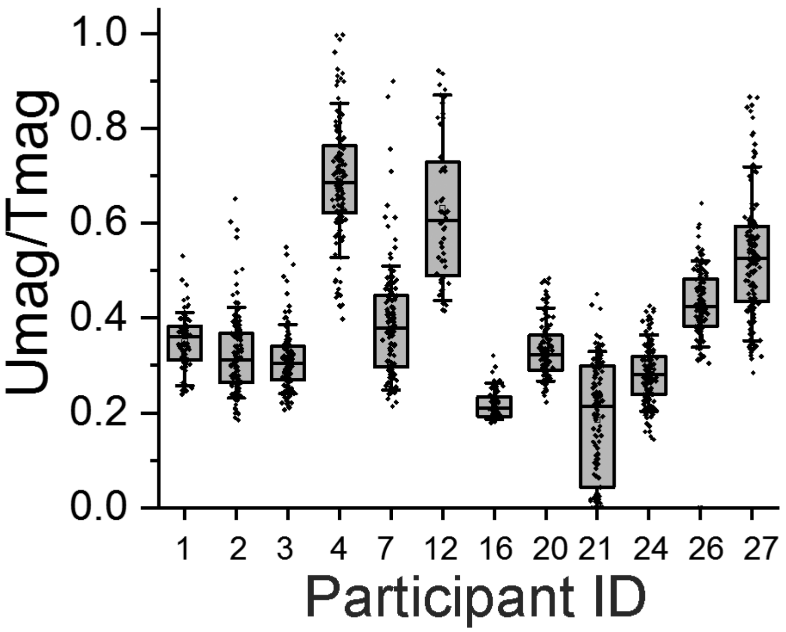

**Supplementary Figure 2**

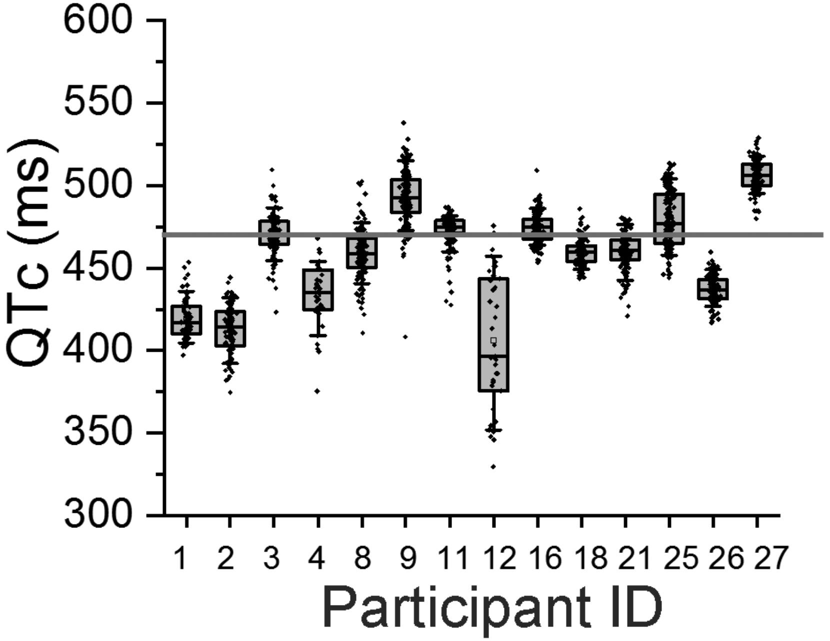

**Supplementary Figure 3**

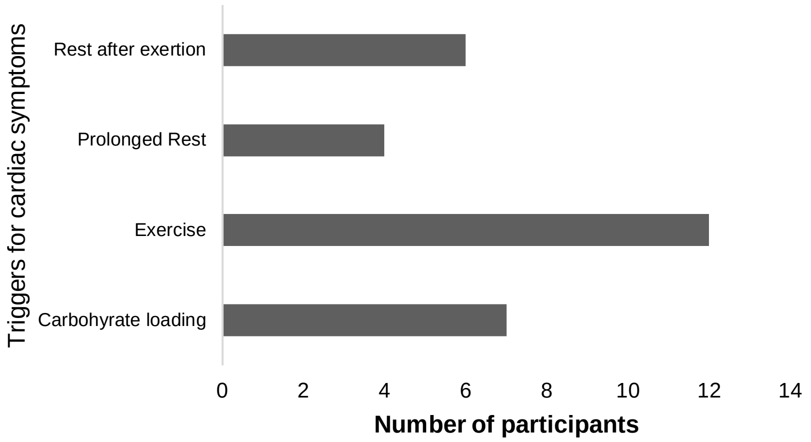
